# BIOMARKERS OF AIRWAY INFLAMMATION AND ASTHMA SYMPTOM CONTROL IN CHILDREN

**DOI:** 10.1101/2023.02.14.23285821

**Authors:** J.S. Alieva, E.G. Furman, E.A. Khuzina, S.T. Abdrakhmanova, M.S. Ponomareva, V.S. Sheludko, V.L. Sokolovsky

## Abstract

**INTRODUCTION:** Bronchial asthma (BA) is a heterogeneous pulmonary disease with various phenotypes based on detection of multiple biomarkers. However, most of the biomarkers are still experimental and present limitations for clinical practice.

**OBJECTIVE:** The purpose of this study was to investigate the relationship between the level of the available T2-inflammatory biomarkers in childhood with poor asthma control and features of asthma management.

**MATERIAL AND METHODS:** The study comprised 100 patients aged 5–17 years (median age 13 years) with an established bronchial asthma diagnosis. The level of asthma control of each patient was assessed by the Asthma Control Test (ACT and C-ACT) and the Composite Asthma Severity Index (CASI). T2-inflammatory biomarkers of the mucous membranes of the respiratory tract include total immunoglobulin E (IgE total) levels, peripheral blood eosinophil levels, fractional exhaled nitric oxide (FeNO) and a nasal smear eosinophil count. A measure of association was determined by standard statistical methods for data analysis.

**RESULTS:** Despite the prescribed basic therapy, the majority of children do not achieve adequate asthma symptom control. This research revealed that 43% of patients had at least one or more elevated markers of T2-inflammation. High levels of IgE total, increased levels of blood eosinophils (≥400 cells/µL), as well as high FeNO values (≥ 20 ppb) prevail in children with partially controlled and uncontrolled asthma. The most significant biomarker of poor asthma control in children is the total serum IgE concentration ≥ 100 IU/ml. In addition, a significant positive correlation was found between peripheral blood eosinophil levels and the ACT/C-ACT scores (r=0.287, p=0.0039).

**CONCLUSION:** Allergic asthma in children is typically associated with Th2-lymphocytes predominance and eosinophilic airway inflammation. T2-inflammatory biomarkers may be useful in assessing airway inflammation activity and asthma-control assessment in children.

## INTRODUCTION

Among the diseases of the internal organs, bronchial asthma (BA) is one of the most important issues of current theoretical and practical medicine [1]. BA is a heterogeneous multifactor pulmonary disease, characterized by a variable airway obstruction caused by chronic inflammation with the involvement of respiratory epithelium, congenital immune system and adaptive immunity [2]. In spite of success in studying the etiology, pathogenesis, early diagnostics of BA, achievements in the development of effective drugs and treatment-and-prophylactic schemes, epidemiological researches demonstrate growth of morbidity all over the world [3].

As reported by the The Global Asthma Network, at present about 334 million people suffer from this disease including 14% of children [4,5]. According to the data of experts of Chronic Respiratory Disease Collaborators (GBD), in 2015 the prevalence of bronchial asthma in the world has increased by 12.6% compared with 1990 [6]. Although BA incidence has been growing during the last decades, nowadays the situation seems to stabilize. However, there are evident regional differences with continuous increase in morbidity in the countries with low and average income level. When evaluating the publications of the last five years, a global prevalence of asthma symptoms among children and adolescents is about 10%. A key pathogenetic link in the formation of bronchial asthma is a chronic inflammation.

Among pediatric patients with atopic BA, there is observed an imbalance between immunoregulatory Th1-and Th2-cytokines with prevailing Th2-lymphocyte activity with inflammatory cytokine generation. Activation of Th2-lymphocytes in the airways is a determinative link in BA pathogenesis. Th2-cells through the cytokines produced by them (IL-4, IL-5, IL-6, IL-10, IL-13) induce in B-lymphocytes and plasmocytes the change in synthesis of IgM to that of IgE as well as an increased adhesion of eosinophils, basophiles and mast cells to the vascular endothelium. Hyperreactivity of smooth muscle fibers of bronchi, hyperplasia of airway mucosa along with immunological imbalance between T-helpers 1 (Th1) and T-helpers 2 (Th2) contribute to allergic inflammation maintenance.

It is known that biomarker is a characteristic (biological sign), which is used as an indicator of the status of the body as a whole. For respiratory diseases including BA, biomarkers can be identified in the sputum, bronchoalveolar lavage fluid (BALF), bronchial bioptate, exhaled air and exhaled air condensate, blood and urine. The main biomarkers and substrates of their obtaining are presented in Fig. 1.

**Figure 1.**
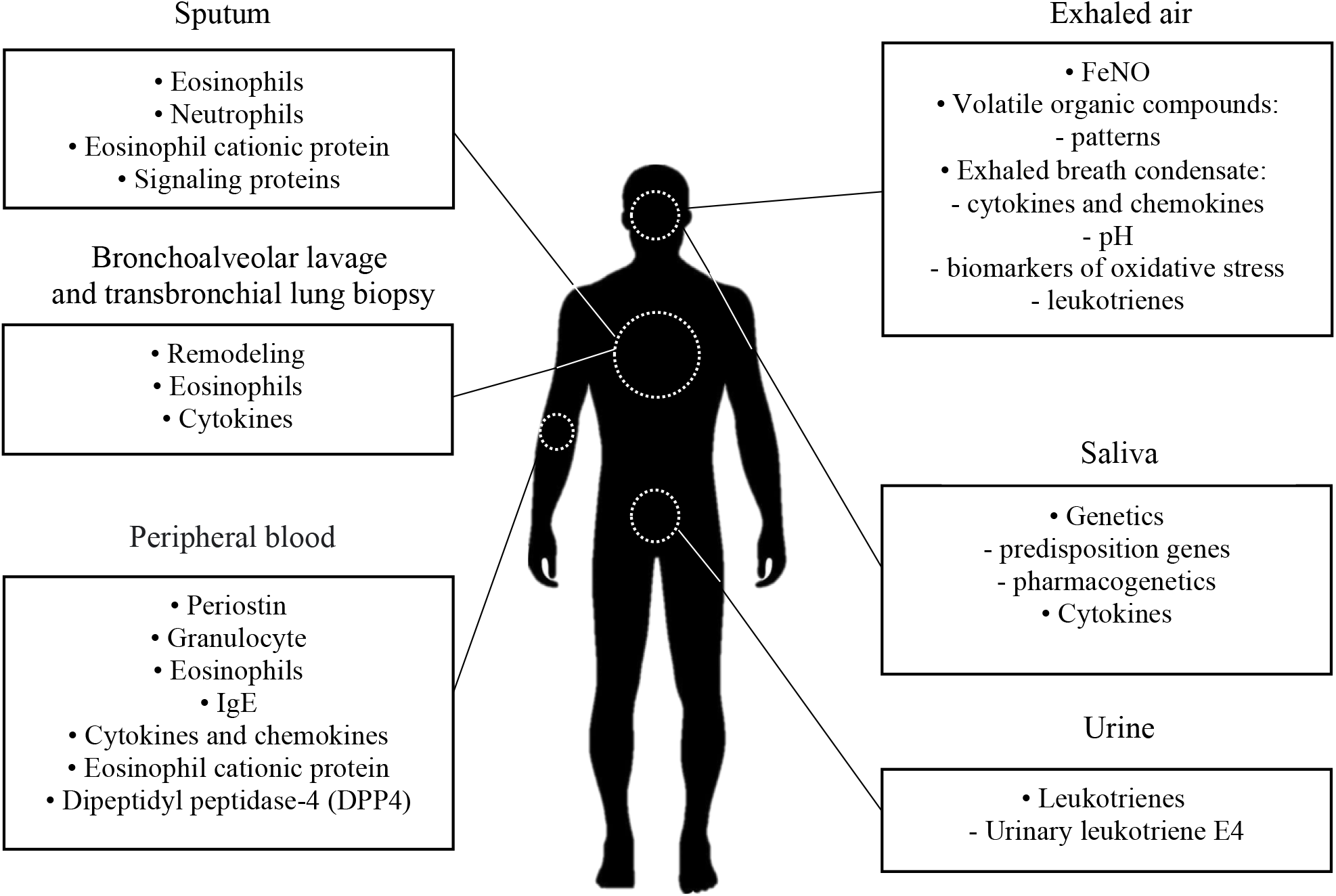
The variability of T2-inflammatory markers [13, 14, 15, 16].

The T2-inflammation of airways, as a rule, is characterized by an increased level of peripheral blood eosinophils ≥ 150 cells/µL in adults (≥ 300 cells/µL in children) and/or ≥ 2 % in the induced sputum as well as by an increased concentration of exhaled air nitric oxide (FeNO ≥ 20 ppb) [8, 10]. Elevation of IgE total content in blood often is accompanied by the development of clinically significant hypersensitivity to allergens [8,10]. Just allergic inflammation with a high level of T2-inflammatory reaction prevails in children with BA compared to adults who have non-allergic inflammation with a low level of signs or with no signs of T2-inflammation. Asthma phenotypes can be used to identify the disease markers, so as to help in diagnostics [12].

Despite a wide availability of inhalation glucocorticosteroids (iGCS) and standardization of recommendations for asthma treatment, the disease control in most children remains nonoptimal. Annually, more than 50% of all BA children have not less than one exacerbation including children with mild asthma. The issue of asthma control is rather relevant in the world since only 20% of patients achieve a complete control of disease [7, 17, 18]. Unfortunately, while the majority of biomarkers applied in BA is used for scientific purposes and for introduction into medical practice, their validization and assessment of diagnostic value is needed [8,9]. However, studying of the airway inflammation markers in patients with BA, especially with an uncontrolled variant, is of great practical significance, since for clinical use, every day immunobiological preparations are registered and administered by indications [10].

## OBJECTIVE

To establish the relationship between the level of biomarkers of airway T2-inflammation in children suffering from BA and the control of disease as well as specificity of its course.

## MATERIAL AND METHODS

In accordance with the principles of the Declaration of Helsinki of the World Medical Association for the period from November 2019 to March 2021, a single-centre prospective cohort study of 100 patients aged 5 to 17 years with the established bronchial asthma diagnosis of different severity and control degree was carried out. Bronchial asthma was diagnosed on the basis of acting clinical recommendations. Before a child was involved in the investigation, an informed written consent from his (her) legal representative in accordance with local laws and regulations was received. The study was carried out on the basis of Regional Children’s Clinical Hospital of Perm and polyclinics of Perm Krai.

Complete sampling method was used to choose the cases. The method for determining the volume of sampling was based on a specialized formula with unknown volume of sampled population.

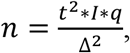

where n-sample volume;

t – confidence coefficient;

I – sampling index (%) taken as 0.5 (with this value, sampling error size is maximum);

q =1-I, a share of respondents without the sign under investigation;

Δ – sampling error limit.

With the confidence level of 95%, the confidence coefficient is equal to 1.96 and the required sampling is assessed as:

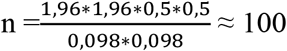

To be included in the research, all the participants underwent complex examination of their clinical condition and assessment of external respiration function. On the basis of the clinical data, peak expiratory rate (PER) parameters, spirography data, there was determined the BA severity criterion using generally accepted criteria [19]. The level of BA control was assessed according to Asthma Control Test – ACT and C-ACT and The Composite Asthma Severity Index – CASI) [3,20]. In addition, all the patients underwent spirography with the assessment of obstruction and identification of the forced expiratory volume per minute (FEV1) and forced vital capacity (FVC) (hand-operated spirometer MicroGP, Great Britain), pulsoxymetry, X-ray examination of the thorax.

All the patients were subjected to collection of disease anamnesis, allergological anamnesis, assessment of relative and absolute peripheral blood eosinophil levels, rhinocytogram, IgE total level in blood serum, exhaled air nitric oxide (FeNo), immunogram by indications. The IgE total concentration in blood serum was investigated using the method of chemiluminescent immune-enzyme analysis. The number of peripheral blood eosinophils was determined using automatic hemoanalyzer; FeNo level was measured according to recommendations of The American Thoracic Respiratory Society using a portative gas analyzer (NObreath, Bedfont Scientific Ltd. BeлиkoбpиTaHия) [21]. The increased markers were considered eosinophil level ≥ 300 cells/µL, FeNO ≥ 20 ppb, IgE total level ≥ 100 IU/ ml [10, 11].

The inclusion criteria: children with established diagnosis of BA age 5 to 17 years (daytime and night-time BA symptoms); decreased functional indicators (FEV1 or PER); presence of exacerbations causing limitation of physical activity and sleep disturbance; absence of contraindications to application of functional and laboratory methods of investigation; signed voluntary informed consent to be involved in research.

The exclusion criteria: any acute respiratory infections for the research period; age not less than 5 years, (due to impossibility to use spirography in this age group); any severe disease or disorder which is able to influence investigation results or patient’s ability to participate in research; patient’s refusal to participate).

Statistical treatment of results was carried out using the statistical program Microsoft Excel 2010. Hypothesis of normality of distribution of the studied parameters was checked using Shapiro-Wilk test.

When comparing the dependent and independent groups of symptoms, characterizing the level of control and/or severity of disease (depending on the type of distributions of analyzed parameters), unpaired Student’s t test or its nonparametric analog – Mann-Whitney U test was used. To analyze the tables of contingency, chi-square criterion was applied. Interconnection of variable data was studied with correlation analysis using Pearson’s linear correlation coefficient (r) and Spearman’s rank correlation coefficient (r). To check the equality of the medians of some samplings, Kraskel-Wallis test was used [23]. Significance level for the examined hypotheses was equal to 0.05. To give a quantitative description of closeness of the ties between two signs, odds ratio (OR) and its 95% confidence intervals were used.

## RESULTS

Characteristic of patients (n=100) involved in research is presented in Table 1. All the children of the examined group were followed up by pulmonologist with the diagnosis of BA. The patient’s age median (Me) was 13 years)Q1–Q3: 9; 15 years). Most patients (72%) were males. 62 patients had mild BA, 27 – moderate BA and 11 children – severe asthma. The majority of children (80%) had an associated allergic rhinitis, 40% of children were diagnosed pollinosis and a third of patients (32%) atopic dermatitis. In 21% of cases, patients were diagnosed degree I-II excess body mass/obesity; in 14% of patients a low level of forced expiratory volume (FEV1) < 70% of the proper level was detected.

**Table 1.** General characteristics of patients with different asthma control. Note: p_1-2-3_ – compared groups according to the asthma control; mTBAs – major tracheobronchial anomalies; BMI – body mass index; ICS – inhaled corticosteroids; FEV1 – forced expiratory volume in the first second; LABAs – long-acting beta-agonists; LTRA – leukotriene receptor antagonists; quantitative data are presented as a median and interquartile range (Q1–Q3), where Q1 is the 25th percentile, Q3 is the 75th percentile; categorical variable are described by absolute and relative frequencies (%); Normality of data distribution was tested with Shapiro-Wilk tests. The Kruskal Wallis test is used to compare more than two independent samples. The p-value 0.05 refers to the significance level in a hypothesis test.

| | Total cohort<br>n=100 | Controlled<br>(n=26) | Partially<br>controlled<br>(n=67) | Uncontrolled<br>(n=7) | $p_{1-2-3}$ |
| --- | --- | --- | --- | --- | --- |
|  |  | 1 | 2 | 3 |  |
| Male/Female gender n (%) | 72 (72) /28(28) | 19 (73) /7(27) | 48 (72) /19(28) | 5 (71) /2(26) | 0,994 |
| Age (y) | 13 (9-15) | 14 (12-16) | 13 (8-15) | 11 (9,5-13,5) | 0,143 |
| Comorbidities, n (%): |  |  |  |  |  |
| allergic rhinitis | 80 (80) | 19 (73) | 57 (85) | 4 (57) | 0,126 |
| atopic dermatitis | 32 (32) | 9 (35) | 21 (31) | 2 (29) | 0,936 |
| hay fever | 40 (40) | 10 (38) | 28 (42) | 2 (29) | 0,756 |
| food allergy | 20 (20) | 7 (27) | 10 (15) | 3 (43) | 0,126 |
| mTBAs | 16 (16) | 4 (15) | 10 (15) | 2 (29) | 0,652 |
| urticaria | 14 (14) | 6 (23) | 7 (27) | 1 (14) | 0,289 |
| Asthma severity, n (%): |  |  |  |  |  |
| mild | 62 (62) | 23 (88) | 38 (57) | 1 (14) | 0,004 |
| moderate | 27 (27) | 2 (8) | 21 (31) | 4 (57) |  |
| severe | 11 (11) | 1 (4) | 8 (12) | 2 (29) |  |
| BMI (kg/m2) | 19,5 (17,1-22,5) | 20,0 (17,3-22,0) | 19,3(16,8-22,6) | 19,0 (17,6-22,0) | 0,976 |
| FEV<sub>1</sub>% | 92,5 (80,0-100,5) | 94,0 (81,3-99,5) | 90,0 (80,0-103,8) | 92,0 (69,0-109,0) | 0,922 |
| Asthma treatment, n (%) |  |  |  |  |  |
| ICS | 35 (35) | 3 (12) | 28 (42) | 4 (57) | 0,011 |
| ICS / LABAs | 30 (30) | 2 (8) | 25 (37) | 3 (43) | 0,015 |
| LTRA | 4 (4) | 0 (0) | 4 (6) | 0 (0) | 0,359 |
| Omalizumab, Mepolizumab | 6 (6) | 1 (4) | 5 (8) | 0 (0) | 0,633 |
| Patients, accessed the emergency department, for the management of exacerbations, n (%) | 34 (34) | 6 (23) | 24 (36) | 4 (57) | 0,207 |
| Number of exacerbations, n | 2,0 (1,0-3,0) | 0 (0-0,8) | 0 (0-1,0) | 1,0 (0-3,0) | 0,071 |

A third of patients (35%) received inhalation glucocorticosteroids (iGCS) as a basic therapy (11% of them in high doses), in 30% of children iGCS was combined with long-acting β2-agonists (LABA). Biological therapy with the preparations of monoclonal antibodies (omalizumab, mepolizumab) ≥ 12 months before inclusion in the research was applied in 6 patients. The rest patients did not receive a regular treatment. 34% of patients, despite the basic therapy carried out, during the previous year had on average two exacerbations of BA. While collecting anamnestic data, the factors of hypoallergenic regime disturbance (domestic animals, carpets, blooming house plants, fungi, passive smoking, gas contamination of street with car exhaust or nearby industrial enterprises) were revealed in 87% of children.

The greatest number of patients (67) had a partial control of BA, in 7 children the absence of control was diagnosed despite the therapy applied; in 26 patients a complete control was achieved.

Among BA patients with uncontrolled course, more often than in children with complete/partial control of disease, there occurred moderate and severe forms of bronchial asthma. In this group high doses of iGCS (p=0.011) were used and more often combined therapy (iGCS/LABA) (p=0.015) was applied. A significant share of patients with uncontrolled BA course (57%) had to ask for emergency care.

The levels of the studied markers and the number of patients with the increased levels are presented in Tables 2 and 3. In 43% of patients under observation, more than one criterion of T2-inflammation was found, at the same time, more often an increased concentration of IgE total in blood serum ≥ 100 IU/ml and the number of blood eosinophils ≥ 300 cells/µL was detected (Fig 2). In the groups of children with partially controlled and uncontrolled course of BA significantly higher IgE total levels (chi-square=7.185, p=0.028), isolated rise in blood eosinophils level ≥400 cells/µL (chi-square = 9.097, p=0.011) as well as FeNO level ≥ 20 ppb (chi-square = 12.000, p=0.003) was noted. In addition, we have established a direct correlation between the absolute blood eosinophil level and the sum of scores ACT/C-ACT (R=0,287, p=0,0039).

**Table 2.** Levels of inflammatory biomarkers in children with different asthma control. Note: p_1-2-3_ – compared groups according to the asthma control; quantitative data are presented as a median and interquartile range (Q1–Q3), where Q1 is the 25th percentile, Q3 is the 75th percentile; categorical variable are described by absolute and relative frequencies (%); Normality of data distribution was tested with Shapiro-Wilk tests. The Kruskal Wallis test is used to compare more than two independent samples. The p-value refers to the significance level in a hypothesis test.

| Biomarkers | Total cohort<br>n=100 | Controlled<br>(n=26) | Partially<br>controlled<br>(n=67) | Uncontrolled<br>(n=7) | p<sub>1-2-3</sub> |
| --- | --- | --- | --- | --- | --- |
|  |  | 1 | 2 | 3 |  |
| IgE total, IU/ml | 182,8 (64,1-481,0) | 132,1 (76,25-613,0) | 167,0 (36,5-464,8) | 254,5 (21,0-663,8) | 0,991 |
| Blood eosinophils,<br>cells/μL | 145,0 (60,25-373,4) | 156,0 (68,05-403,3) | 145,0 (58,8-370,0) | 127,5 (0-445,0) | 0,818 |
| FeNO, ppb | 11,0 (6,0-18,0) | 11,5 (8,5-31,0) | 10,0 (3,0-13,5) | 15,0 (13,0-17,0) | 0,275 |

**Table 3.** Characteristics of the various levels of inflammatory biomarkers in children with different asthma control. Note: The χ2-test was used to compare categorical variables. The p-value 0.05 refers to the significance level in a hypothesis test.

|  | Total<br>cohort<br>n=100 | Controlled<br>(n=26) | Partially<br>controlled<br>(n=67) | Uncontrolled<br>(n=7) | p<sub>1-2-3</sub> |
| --- | --- | --- | --- | --- | --- |
|  |  | 1 | 2 | 3 |  |
| Patients n (%) showed IgE total levels $>100$ IU/mL | 43 (43) | 10 (39) | 29 (43) | 4 (57) | 0,028 |
| Patients n (%) showed blood eosinophils levels |  |  |  |  |  |
| ≥ 300 cells/μL | 24 (24) | 7 (27) | 15 (22) | 2 (29) | 0,509 |
| ≥ 400 cells/μL | 14 (14) | 5 (19) | 8 (31) | 1 (14) | 0,011 |
| ≥ 500 cells/μL | 11 (11) | 3 (12) | 7 (10) | 1 (14) | 0,685 |
| Patients n (%) showed FeNO levels |  |  |  |  |  |
| ≥20 ppb | 8 (8) | 3 (12) | 5 (8) | 0 (0) | 0,003 |
| ≥50 ppb | 6 (6) | 1 (4) | 5 (8) | 0 (0) | 0,016 |
| ≥100 ppb | 2 (2) | 0 (0) | 2 (3) | 0 (0) | 0,049 |

**Figure 2.**
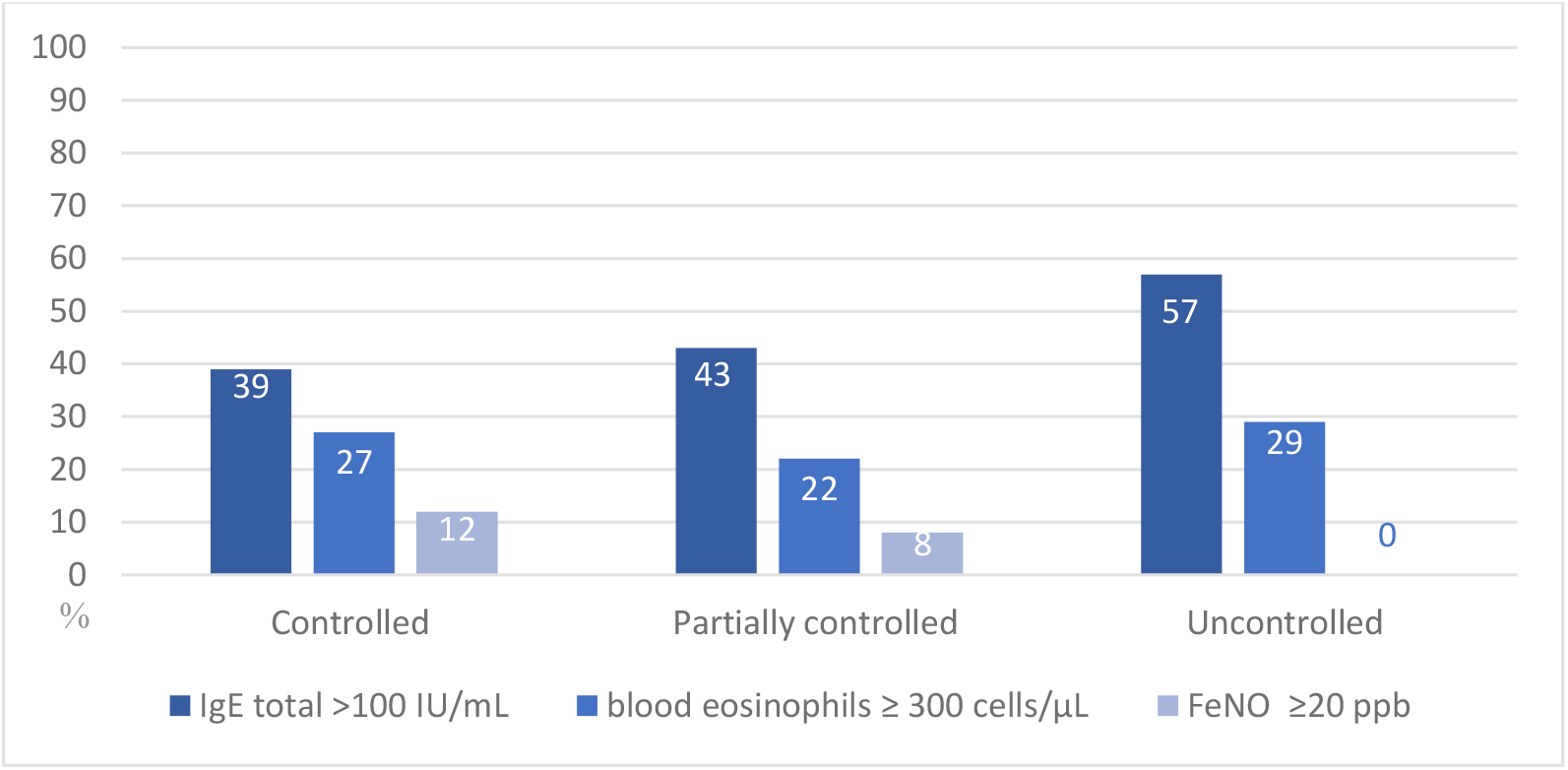
T2-inflammatory biomarkers in patients with different asthma control; %;

According to the results of analysis of odds ratio (OR) of control unattainability, the most significant inflammation biomarkers influencing achievement of controlled BA course were found (Table 4).

**Table 4.** T2-inflammatory biomarkers in the absence of adequate asthma control in children.

| Biomarkers | OR | Confidence interval (CI) (95%) |
| --- | --- | --- |
| IgE total, IU/ml | 1,280 | 0,73-2,25 |
| Blood eosinophils, cells/μL | 0,808 | 0,43-1,53 |
| FeNO, ppb | 0,552 | 0,21-1,47 |

## DISCUSSION

The most significant biomarker with the absence of optimal control of bronchial asthma in children is an increased concentration of IgE total ≥ 100 UI/ml (OR=1,280; CI [0,73-2,25]). Thus, elevation of IgE total level in blood serum by more than 20% increases the risk of partially controlled and uncontrolled course of BA.

It is impossible to extrapolate the dependence of the degree of BA control on the level of eosinophil inflammation biomarkers among children to the child population as a whole, due to the small number of samples under investigation. Perhaps, while increasing the sampling size the results will differ from those presented in this article.

In the group of children under examination, the majority (74%) does not achieve an optimal level of control of disease in spite of the basic therapy applied. However, it is worth noting that not all the children received a rational basic therapy, and the share of those, who regularly applied inhaled corticosteroids, was only 30%. Among children, an allergic phenotype of BA proceeds with the dominating activity of Th2-lymphocytes and development of airway eosinophil inflammation. The most frequent markers of allergic inflammation are increased levels of peripheral blood eosinophils and increased IgE total concentration in blood serum. The most important biomarkers, negatively influencing achievement of the controlled course of BA, are high IgE total levels ≥ 100 IU / ml, increased blood eosinophil level ≥ 400 cells/µL as well as FeNO level ≥ 20 ppb.

Further studying of relationship between biomarkers and clinical features of BA as well as development of early and individual choice of significant biomarkers can contribute to prevention of nonreversible changes in the bronchial wall (remodeling), reduction of drug load and improvement of diagnostics and prognosis for BA course in children.

Gratitude.

This research was supported by a joint grant from the Israeli Ministry of Science and Technology (MOST, 3-16500) from the Russian Foundation for Basic Research (RFBR) (joint research project 19-515-06001).

## Data Availability

All data produced in the present study are available upon reasonable request to the authors

## REFERENCES

1. GINA Report, Global Strategy for Asthma Management and Prevention. 2015

2. Borta, S.M.; Dumitra, S.; Miklos, I.; Popetiu, R.; Pilat, L.; Puşchiţ ă, M.; Marian, C. Clinical Relevance of Plasma Concentrations of MBL in Accordance with ige Levels in Children Diagnosed with Bronchial Asthma. Medicina 2020, 56, 594. Https://doi.org/10.3390/medicina56110594

3. National program “Bronchial asthma in children. Treatment strategy and prevention”. 3rd ed., rev. And extra. - M.: Publishing house “Atmosphere”, 2008 (In Russian).

4. Network GA. The Global Asthma Report, Auckland, New Zealand. (2018).

5. Avdeev S.N., Nenasheva N.M., Zhudenkov K.V. and others. Prevalence, incidence, phenotypes and other characteristics of severe bronchial asthma in the Russian Federation // Pulmonology. 2018. V. 28. No. 3. S. 341–358 (In Russian).

6. GBD 2015 Chronic Respiratory Disease Collaborators. Global, regional, and national deaths, prevalence, disability-adjusted life years, and years lived with disability for chronic obstructive pulmonary disease and asthma, 1990-2015: a systematic analysis for the Global Burden of Disease Study 2015 // Lancet Respir. Med. 2017. Vol. 5. ? 9. P. 691–706.

7. Allegra L, Cremonesi G, Girbino G, et al. Real-life prospective study on asthma control in Italy: cross-sectional phase results. Respir. Med. 2012; 106 (2): 205–214. https://doi.org/10.1016/j.rmed.2011.10.001

8. Ненашева Н.М. Значение биомаркеров в диагностике и терапии бронхиальной астмы// Практическая пульмонология. 2017. №4.

9. Vijverberg SJ, Hilvering B, Raaijmakers JA, Lammers JW, Maitland-van der Zee AH, Koenderman L. Clinical utility of asthma biomarkers: from bench to bedside. Biologics. 2013;7:199–210.

10. Pampura A.N., Kamaev A.V., Lebedenko A.A. Biomarkers of asthma in children. New opportunities, real practice and prospects. Medical Bulletin of the South of Russia. 2022;13(2):91-101. (In Russian).

11. Konradsen J.R., Skantz E., Nordlund B., Lidegran M., James A. et al. Predicting asthma morbidity in children using proposed markers of Th2-type inflammation. Pediatr Allergy Immunol. 2015; 26:772–779.

12. Udoko M., De Keyser H., Szefler S. J. Should children with asthma simply be treated as little adults? //Annals of allergy, asthma & immunology: official publication of the American College of Allergy, Asthma, & Immunology. – 2021. – T. 127. – No. 5. – C. 520-521

13. Breiteneder H, Peng YQ, Agache I, Diamant Z, Eiwegger T, Fokkens WJ, Traidl-Hoffmann C, Nadeau K, O’Hehir RE, O’Mahony L, Pfaar O, Torres MJ, Wang DY, Zhang L, Akdis CA. Biomarkers for diagnosis and prediction of therapy responses in allergic diseases and asthma. Allergy. 2020 Dec;75(12):3039–3068.

14. Tiotiu A. Biomarkers in asthma: state of the art. Asthma Res Pract. 2018 Dec 21;4:10.

15. Guida G, Bagnasco D, Carriero V, Bertolini F, Ricciardolo FLM, Nicola S, Brussino L, Nappi E, Paoletti G, Canonica GW, Heffler E. Critical evaluation of asthma biomarkers in clinical practice. Front Med (Lausanne). 2022 Oct 10;9:969243. doi: 10.3389/fmed.2022.969243.

16. Porsbjerg C, Melén E, Lehtimäki L, Shaw D. Asthma. Lancet. 2023 Jan 19:S0140–6736(22)02125-0. doi: 10.1016/S0140-6736(22)02125-0.

17. Basharat S, Jabeen U, Zeeshan F, Bano I, Bari A, Rathore AW. Adherence to asthma treatment and their association with asthma control in children. J Pak Med Assoc. 2018 May;68(5):725–728.

18. Haughney J, Price D, Kaplan A, Chrystyn H, Horne R, May N, Moffat M, Versnel J, Shanahan ER, Hillyer EV, Tunsäter A, Bjermer L. Achieving asthma control in practice: understanding the reasons for poor control. Respir Med. 2008 Dec;102(12):1681–93. doi: 10.1016/j.rmed.2008.08.003. Epub 2008 Sep 23. PMID: 18815019.

19. Global Initiative for Asthma. Global strategy for asthma management and prevention. Available from http://www.ginasthma.org. Date last updated, 2022.

20. Wildfire JJ, Gergen PJ, Sorkness CA, Mitchell HE, Calatroni A, Kattan M, Szefler SJ, Teach SJ, Bloomberg GR, Wood RA, Liu AH, Pongracic JA, Chmiel JF, Conroy K, Rivera-Sanchez Y, Busse WW, Morgan WJ. Development and validation of the Composite Asthma Severity Index-an outcome measure for use in children and adolescents. J Allergy Clin Immunol. 2012 Mar;129(3):694–701. doi: 10.1016/j.jaci.2011.12.962. Epub 2012 Jan 12. PMID: 22244599; PMCID: PMC3294274.

21. Khatri SB, Iaccarino JM, Barochia A, Soghier I, Akuthota P, Brady A, Covar RA, Debley JS, Diamant Z, Fitzpatrick AM, Kaminsky DA, Kenyon NJ, Khurana S, Lipworth BJ, McCarthy K, Peters M, Que LG, Ross KR, Schneider-Futschik EK, Sorkness CA, Hallstrand TS; American Thoracic Society Assembly on Allergy, Immunology, and Inflammation. Use of Fractional Exhaled Nitric Oxide to Guide the Treatment of Asthma: An Official American Thoracic Society Clinical Practice Guideline. Am J Respir Crit Care Med. 2021 Nov 15;204(10):e97–e109. doi: 10.1164/rccm.202109-2093ST. PMID: 34779751; PMCID: PMC8759314.

22. 2Lemeshko B. Yu., Lemeshko S. B. Comparative analysis of criteria for checking the distribution deviation from the normal law // Metrology. - 2005. - T. 2. - S. 3-23. (In Russian).

23. Grzhibovsky A. M., Ivanov S. V., Gorbatova M. A. Comparison of quantitative data of three or more independent samples using Statistica and SPSS software: parametric and non-parametric criteria // Science and Health. – 2016. – no. 4. - S. 5-37 (In Russian).

